# AI portal tract detection and characterisation for a regional analysis of steatosis and inflammation in MASLD, MASH, and AIH

**DOI:** 10.1101/2025.04.23.25326290

**Authors:** Dylan Windell, Alastair Magness, Cayden Beyer, Helena Thomaides Brears, Sarah Larkin, Kezia Hobson, Paul Aljabar, Kenneth Fleming, Eve Fryer, Timothy Kendall, Reema Kainth, Phil Wakefield, Caitlin Langford, Pierre Bedossa, Robert Goldin

## Abstract

**Background & Aims:** Annotation of liver biopsies, for disease staging is increasingly aided by digital pathology, however existing systems do not quantify inflammation and steatosis within an anatomical framework. We developed an AI system to quantify portal tracts (PT) and disease features and their regional distribution in Metabolic Dysfunction-Associated Steatotic Liver Disease (MASLD)/Metabolic Dysfunction-Associated Steatohepatitis (MASH) and Autoimmune Hepatitis (AIH).

**Methods:** Digitised images of haematoxylin and eosin-stained specimens were pooled from 4 clinical cohorts (n=390: 89 MASLD, 238 MASH, 63 AIH). Portal tracts, regional steatosis, interface hepatitis, portal and lobular inflammation were quantified using a proprietary AI system and scored by expert pathologists.

**Results:** The percentage of steatosis was higher in MASH (7.5%) compared to MASLD (3.2%, p<0.0001). Lobular regions had larger steatotic vesicles (260 vs 190mm^2^, p<0.0001). AI-derived steatosis quantification correlated with manual grading (r_s_=0.72; p<0.0001). The inflammatory cell number (ICN) was 2-fold higher in AIH compared to MASLD/MASH in interface [390 vs 140; p<0.0001], portal [4600 vs 1500], and lobular [1500 vs 650] regions. Severity of portal inflammation from manual grading correlated well with ICN count at PT (r_s_=0.71; p<0.0001) but not lobular regions (r_s_=<0.1). For equivalent grades of portal inflammation, the ICN was up to 3-fold higher in AIH than in MASLD/MASH (r_s_=0.71; p<0.0001).

**Conclusion:** Despite equivalent pathologist portal inflammation grades, the digital burden of inflammation was significantly higher in AIH than MASLD/MASH. This digital system quantifies PT, inflammation, and steatosis, providing powerful decision support for pathologists in AIH diagnosis and MASH staging.

**LAY SUMMARY:** The detection of portal tracts is an important part of liver biopsy sample quality control and histological scoring. Using artificial intelligence, this study demonstrates a system that automatically detects and quantifies portal tracts and surrounding patterns of inflammation and steatosis. AI found that inflammation was in similar regions but was higher in autoimmune hepatitis than in metabolic dysfunction-associated steatohepatitis, despite similar grading from manual scoring. This AI system provides granular information that can aid biopsy grading and provide insights into liver disease progression and diagnosis.

## Introduction

Metabolic dysfunction-associated steatotic liver disease (MASLD) affects one third of the global population^1^ and is characterised by increased fat in and around hepatocytes^2^. Progression to steatohepatitis (MASH), with concurrent liver inflammation, hepatocyte ballooning, and increased risk of fibrosis, is estimated to occur in 5-14% of adults and projected to increase^1,3,4^. Autoimmune hepatitis (AIH) is rarer, prevalent in 16 cases per 100,000 people globally^5^, but progresses rapidly once diagnosed. Liver biopsy remains the reference standard for diagnosing AIH and staging MASLD/MASH, particularly in complex cases^2,6–8^. Therapeutics for AIH are relatively well established but the metabolic comorbidities and multiplicity of disease features in MASH complicate drug efficacy and have accelerated development of anti-steatotic^9^, appetite suppressing^10^, anti-inflammatory^11^, anti-apoptotic^12^, and anti-fibrotic^13^ candidates. Resolution of histological disease features are primary endpoints in MASH late-phase clinical trials. While clearly improved by blinded and centralised liver histology reading, both inter– and intra-rater variability still affect biopsy-based evaluation^14–17^. This highlights a continued need for standardisation of biopsy sampling and consensus in grading^18^.

Identification of portal tracts (PT) in liver biopsy samples allows the assessment of biopsy adequacy and staging of disease based on the relative anatomical distribution of liver disease features^19–21^. Identification of PT is challenging because they vary greatly in their size, structure, and shape^20^, and their appearance is affected by biopsy needle size, cutting angle^22,23^, and disease stage^24–26^. In recent audits^20,27^, at least 50% of biopsies fail adequacy criteria, so improving PT detection is also desirable.

Artificial Intelligence (AI)-based systems have been developed that complement manual annotation and pathology grading of liver biopsy samples^28^. Existing systems for detecting and segmenting PT have been effective and accurate, with examples being considered by the FDA for application in MASH clinical trials^29^. Automated systems can show high degrees of correlation between manual annotations and automatic measures^30–36^, and are increasingly able to distinguish and segment anatomical features such as interface hepatitis and bile ducts^37^. However, current systems have been developed for fibrosis, and this prevents identification of inflammation, ballooning, and steatosis, which require a different staining agent (Haematoxylin & Eosin, H&E, rather than Masson’s Trichrome or Picrosirius Red). Furthermore, despite the accuracy of these systems, they lack an anatomical framework to assist with interpretation and grading of pathology.

We set out to address these shortcomings by creating an AI system capable of detecting PT and quantifying pathological features relative to PT and other landmarks from biopsy samples stained with H&E. Using this new method, we aimed to comprehensively evaluate the burden of steatosis and of inflammation in a large dataset of MASLD/MASH and AIH liver biopsies. Additional objectives were to examine regional distributions of these features, at PT and in liver lobules, and associations to disease severity from manual grading.

## Materials and Methods

### Study Design and Populations

Data was obtained from four independent clinical cohorts detailed previously^38–40^. Inclusion of digitized datasets was based on adequacy of biopsy samples^41,42^, in terms of stain and sample quality, and on confirmation of AIH^43^ and MASLD/MASH based on clinical criteria and pathology scoring independent of this work^41,42,44^ (**Supplementary Figure 1**). Participants with MASLD/MASH or AIH were enrolled at routine patient visits in secondary or tertiary care, at sites in the United Kingdom, the United States, Poland, and Japan. Patients with any other known chronic liver disease were excluded from this analysis. Where biopsies from multiple timepoints were available, only the baseline timepoint was selected to avoid duplication. All patients and volunteers provided written informed consent. All parent studies were conducted in accordance with the ethical principles of the Declaration of Helsinki 2013 and the Good Clinical Practice guidelines and all studies approved by ethical committees (Institutional review at trial sites for NCT03551522, Ethics committee at Yokohama City Hospital for UMIN000026145, West Midlands – Black Country Research Ethics Committee 14/WM/0010 for ISRCTN39463479, Komisja Bioetyczna przy Instytucie Pomnik-Centrum Zdrowia Dziecka 11/KBE/2016 for NCT03198104).

### Pathology Protocol

*Histological scoring* Histological WSI were split between four pathologists with over 100 years of combined experience, blinded to all clinical data (EF, KF, TK & RG, **Supplementary Table 5**). Biopsy sample adequacy was assessed using the definition outlined by the Royal College of Pathologists^45^.

All MASLD/MASH and AIH WSI were scored using the Ishak scoring system^46^ for portal, interface and lobular inflammation and the NAS scoring system^47^ for lobular inflammation, fibrosis and steatosis.

### AI system for portal tract detection and quantification of steatosis and inflammation

We developed a proprietary system of interrelated AI systems from H&E images that could identify landmark features, detect PT, perform regional demarcation, and spatial analysis to quantify steatotic vesicles and inflammatory cells (**Figure 1**). The following measurements were derived within portal, interface, periportal, and lobular regions:

**Figure 1.**
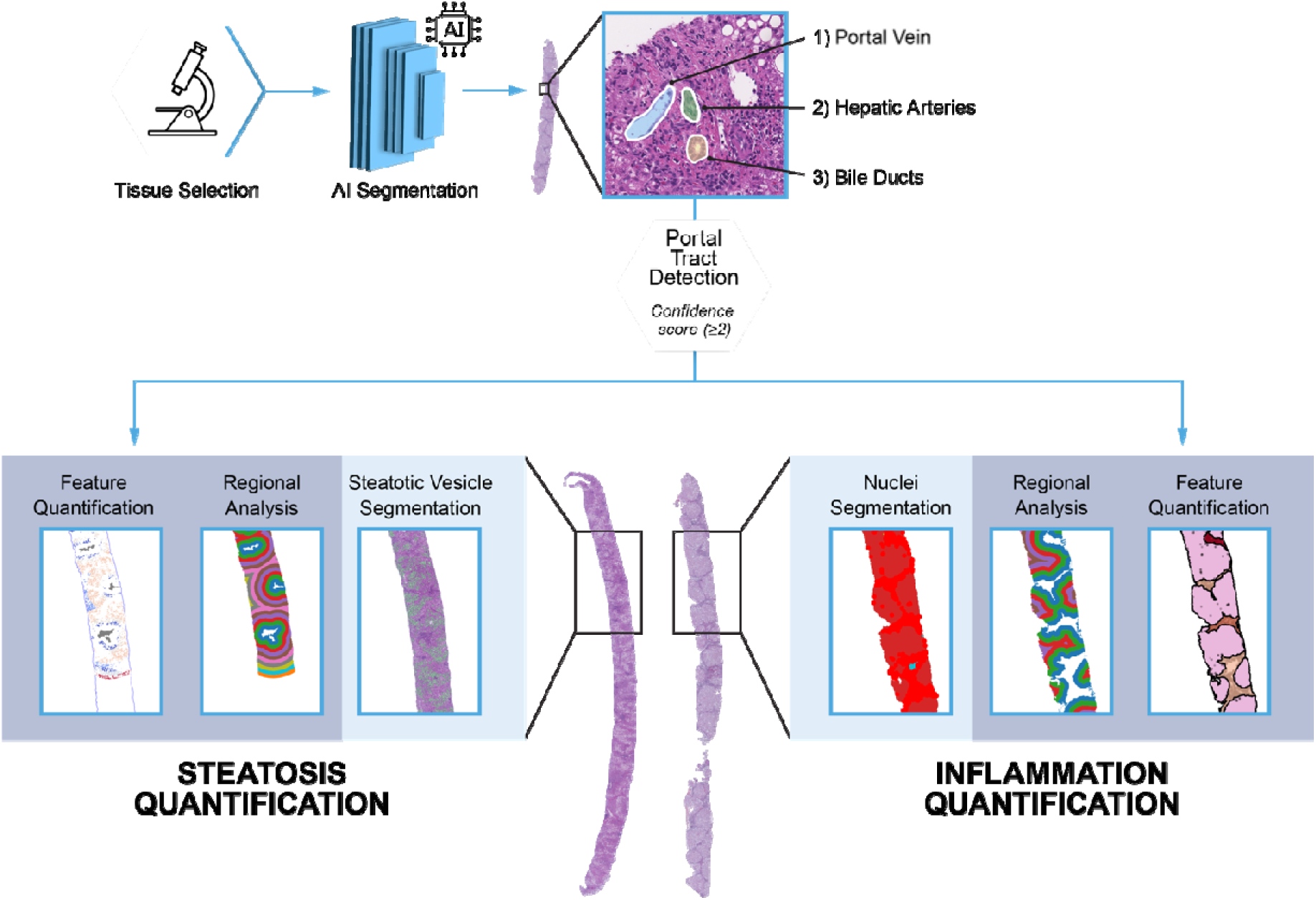
– AI pipeline to segment inflammation and steatosis in relation to portal tract location. Equidistant regions (100µm) from PT allow for detailed quantification of steatosis and inflammation in portal, periportal and lobular regions of liver samples.

Steatosis Fraction (SF) = Area of steatosis / Area of region

Vesicle Hepatocyte Fraction (VHF) = Total number of steatotic vesicles / Total number of hepatocytes

Inflammatory Cell Number (ICN) = Total number of Inflammatory cells

Inflammatory Burden (IB) = Number of inflammatory cells / Area of region

### Statistical analyses

Descriptive statistics were used to summarize cohort characteristics. Median and IQR (Interquartile Range) were used to describe all non-normally distributed variables. Frequency and percentage were used to describe categorical variables.

Differences between AI measurements across disease states were assessed using Wilcoxon rank-sum test. Correlations between AI measurements and pathologist scoring were assessed using Spearman’s rank correlation. Grouped scores between pathologists were either compared using Kruskal Wallace tests or, if there were less than 3 scores within each category, a Wilcoxon rank-sum test was performed. Correlations between AI measurements in different regions were assessed using Pearson’s correlation coefficient.

All statistical analyses were performed in R Studio version 4.2.2. Values of p<0.05 were considered statistically significant.

## Results

### Study populations

A total of 390 individuals were recruited from secondary care and paediatric clinics between February 2014 and March 2019 (**Supplementary Figure 1**). Their mean age was 51 years, 48% were male, with mean BMI of 29.5kg/m^2^ (**Supplementary Table 1**). Overall, 89/390 (23%, all adults) had been diagnosed with MASLD, 238/390 (61%, all adults) with MASH and 63/390 (16%, of which 3% adults) had AIH. In those with MASLD/MASH, known comorbidities were obesity (51%), diabetes (35%), hypertension (21%) and dyslipidaemia (33%). These comorbidities were rare in those with AIH. Treatment for AIH (56%) was considered as ongoing with either Steroids or Azathioprinum.

In those with MASH, 286 had “at-risk MASH” (NAS activity score ≥4 and ≥2 fibrosis stage). Similarly, for AIH disease, 7 had moderate-severe disease (≥2 lobular and portal inflammation score).

Every individual underwent a liver biopsy. Biopsy samples were scored manually by 1-4 pathologists and quantified with the AI-based quantification system of PT detection and liver tissue characterisation described previously (**Figure 1**).

### Portal tract quantification using AI

The AI system identified a total of 7359 PT across the 390 digitized whole slide images (WSI), with a mean number of 19 PT per WSI and 7 PT per 10mm of liver sampled (**Supplementary Table 2**). 378/390 (97%) WSI met the minimum 6-11 PT/sample and 325/390 (83%) WSI met the minimum 10-12 PT/sample quality criterion recommended for pathological assessment^21^. The PT count correlated with length of sample (Spearman’s rho 0.71, p<0.0001) (**Supplementary Figure 2**).

PT detection by AI was comparable in performance with manual detection by each pathologist (**Supplementary Table 3**). Agreement in PT detection by different pathologists ranged in F1 score from 0.58 ±0.16 to 0.72 ±0.11. Whereas for AI, the F1 score was 0.66 ±0.18 against the consensus detection achieved by all pathologists.

### Quantification of steatosis in MASLD and MASH

The AI system delineated 1,658,463 steatotic vesicles from the subset of 327 WSI with MASLD or MASH, with a mean 3732 vesicles per sample for MASLD cases and 5572 vesicles per sample for MASH (**Supplementary Table 2**).

Across the whole of the tissue sampled, the % area with steatotic vesicles (steatosis fraction, SF) was lower in MASLD cases compared to the SF in people with MASH (3.2% (IQR 5.6) vs 7.5% (IQR 7.1), p<0.0001) (**Figure 2, Supplementary Figure 3**). Similarly, the ratio of vesicles to hepatocytes (the vesicle hepatocyte fraction) was lower in MASLD (10.2%; IQR 12.1) than in MASH (20.1% IQR 17.2). The SF correlated with the vesicle hepatocyte fraction (Spearman’s rho=0.9; p<0.0001). The size of steatotic vesicles also differed. Median area of steatotic vesicles was 190μm^2^ (IQR 110) in those with MASLD and 260μm^2^ (IQR 100) in MASH (p<0.0001) (**Figure 3**).

**Figure 2.**
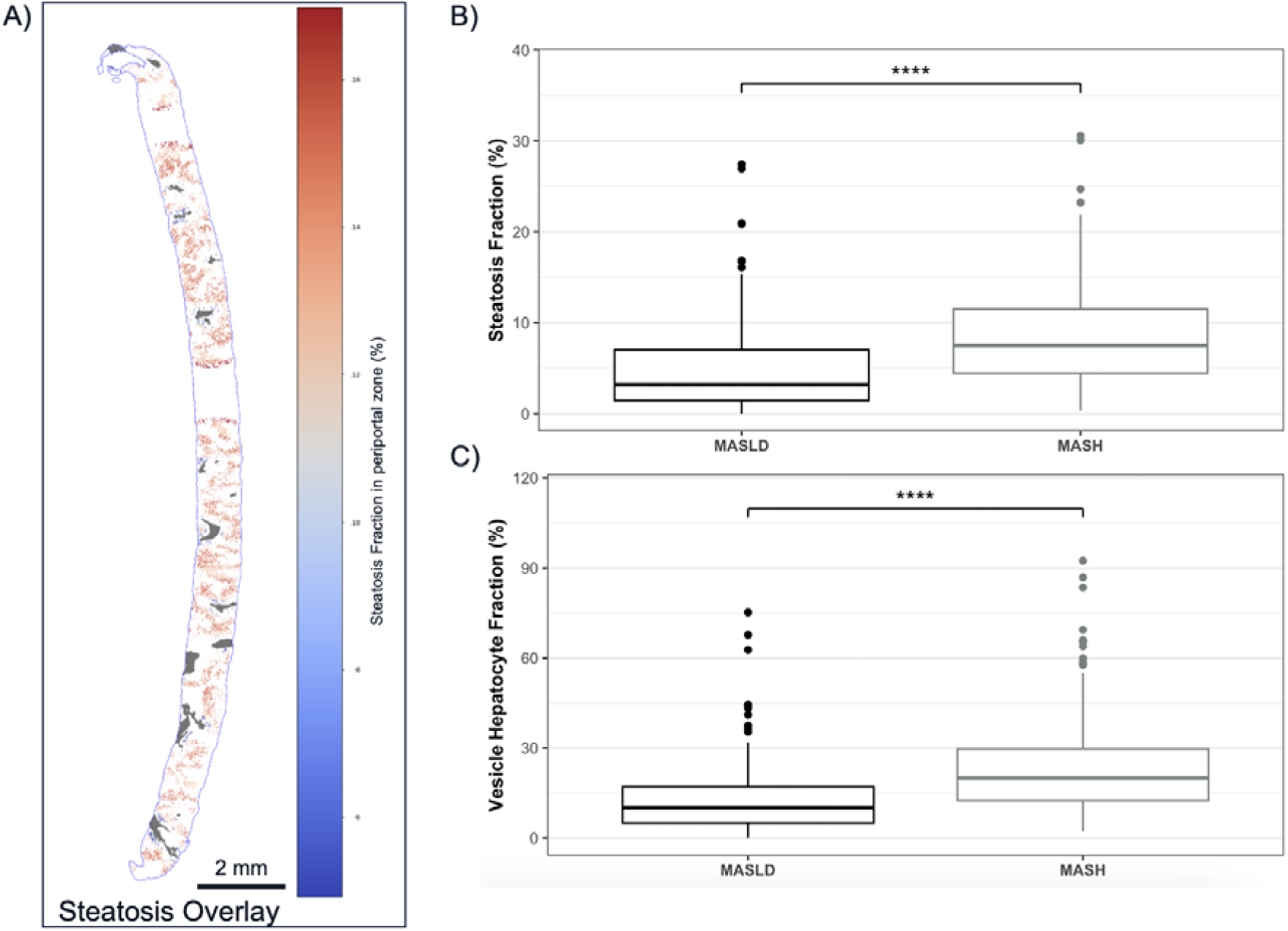
– Steatosis in MASLD (n=89) and MASH (n=238). (A) Example case with an interactive steatosis overlay of steatosis fraction represented on a colorimetric scale (blue-red) at specific regions from PT (grey). (B) Steatosis fraction (%) in lobular regions in MASLD and MASH. (C) Vesicle to hepatocyte fraction (%) in lobular regions in MASLD and MASH.

**Figure 3.**
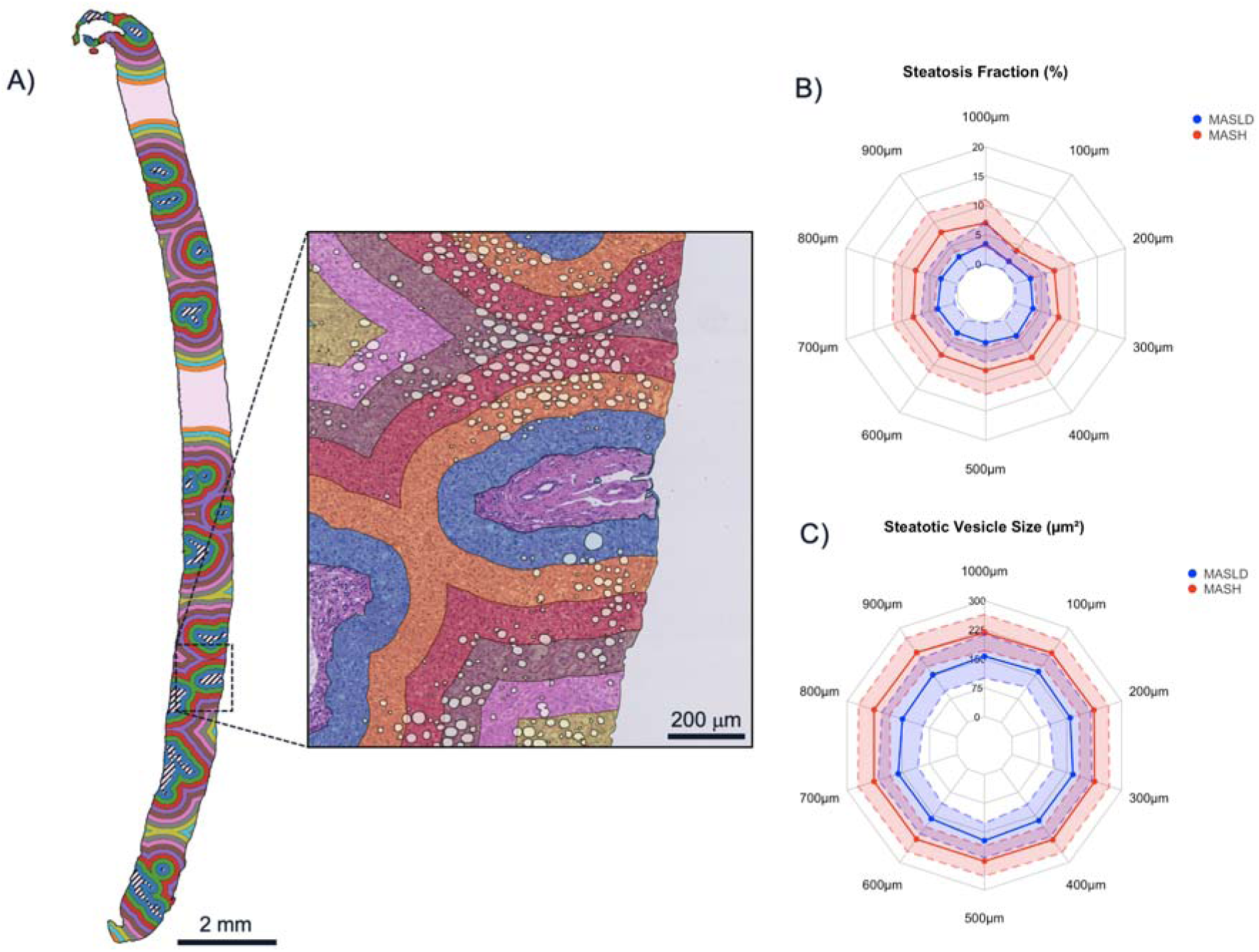
– Regional analysis of steatosis in MASLD (n=89) and MASH (n=238). (A) Steatotic vesicles were quantified within areas defined by regions positioned equidistant from the PT, at intervals of 100μm, in a case with MASLD. (B) Steatosis fraction (%) in MASLD and MASH. (C) Size (µm^2^) of steatotic vesicles in MASLD and MASH. Median is represented as a solid line and the upper/lower interquartile range is represented as a dashed line.

#### Regional distribution of steatosis

In the immediate vicinity of the PT, up to 100μm from the PT, the SF was lower compared to further away in lobular regions, with higher levels of steatosis observed at 500μm from the portal boundary (**Figure 3**). This was observed for both MASLD and MASH. Significant differences were measured in all regions measured up to 1mm from each PT. The biggest difference in steatotic vesicle aggregation was at approximately 900μm from the PT, with SF of 2.7% (IQR 5.9) for MASLD and 7.9% (IQR 4.2) for MASH (p<0.0001). The difference in vesicle size between MASLD and MASH was preserved at all distances from the PT.

#### Association with steatosis grading from manual pathology reads

Expert pathologist grading for steatosis did not differ significantly between MASLD and MASH (**Supplementary Figure 3**). Both AI-derived metrics for steatosis quantification showed significant correlation with steatosis grading in lobular regions in both MASLD (for SF r_s_=0.61 and for VHF r_s_=0.64; p<0.0001] and MASH (for SF r_s_=0.7 and for VHF r_s_=0.72; p<0.0001] across all 3 pathologists together or individually.

### Quantification of inflammation in MASLD/MASH and AIH

The AI system identified 1.5 million inflammatory cells at PT and 0.4 million inflammatory cells in lobular regions, and 0.11 million cells in interface regions across all 390 WSI.

Inflammatory cells were detected in the liver tissue of both MASLD or MASH individuals (ICN: 4300 mean number of cells per WSI) and in those with AIH (ICN: 12000 mean number of cells per WSI) (**Supplementary Table 2**). The inflammatory burden (IB) was higher in AIH compared to MASLD/MASH at PT (2700 cells/mm^2^ (IQR 2800), vs 1400 cells/mm^2^ (IQR 1800), p<0.0001) and in lobular regions (60 cells/mm^2^; IQR 90 vs 40 cells/mm^2^; IQR 50, p<0.001) (**Figure 4**).

**Figure 4.**
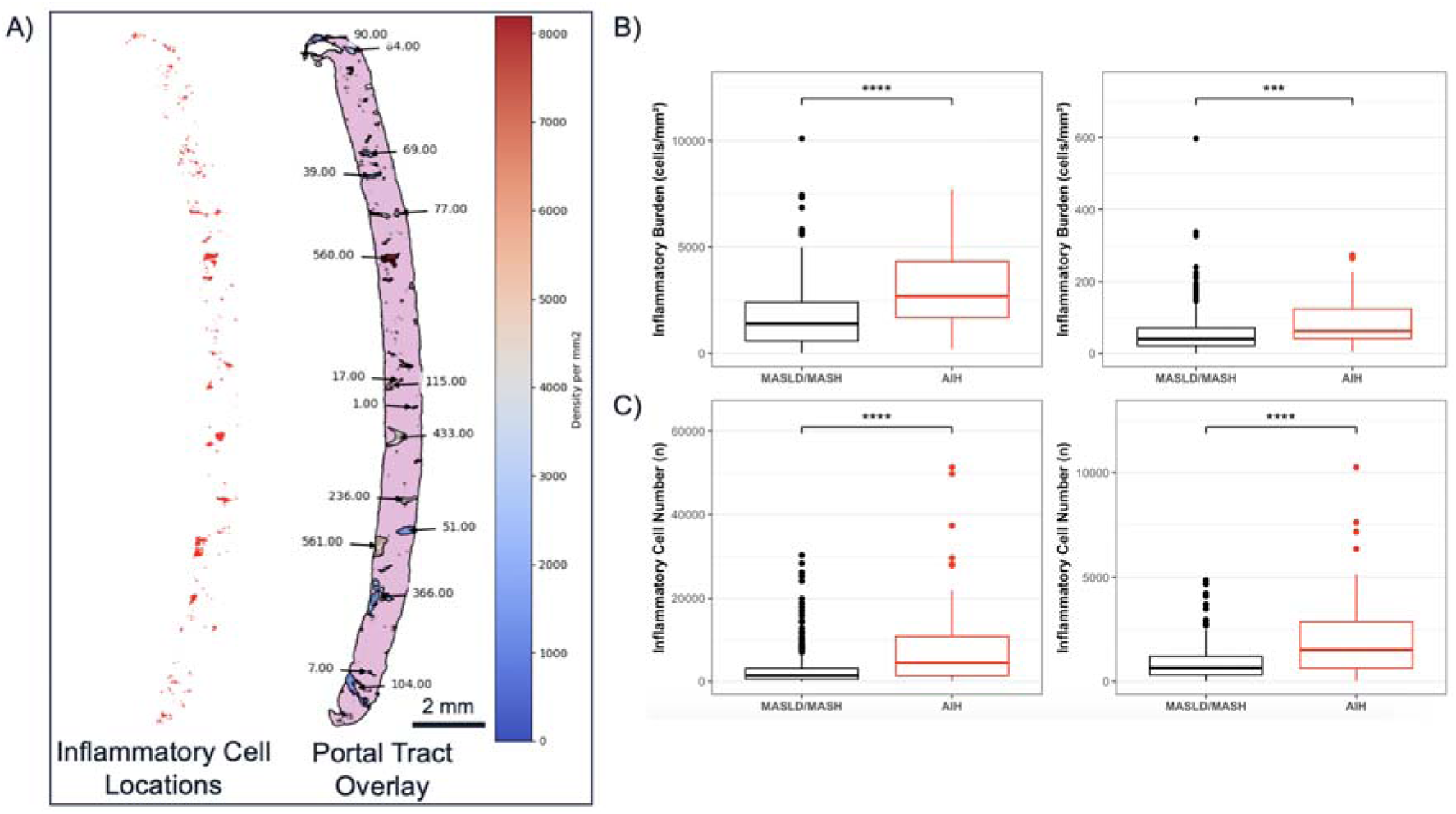
– Comparison of inflammation in MASLD/MASH (n=327) versus AIH (n=63). (A) Example of a case with MASLD; both segmented inflammatory cells (ICN) and inflammatory burden (IB) were quantified and overlayed with the anatomical location of the PT and represented with a colorimetric scale. (B) The IB (cells/mm^2^) and (C) ICN in portal and lobular regions in MASLD/MASH and AIH.

#### Regional distribution of inflammation

There was significantly more inflammation in all regions extending from PT in AIH cases compared to MASLD/MASH, but the regional distribution was similar in both diseases. Maximal ICN and IB were in the PT for both AIH and MASLD/MASH (**Supplementary Table 2, Figure 5**). At the immediate vicinity of the PT, up to 100μm from the PT, IB was suggestive of interface hepatitis in AIH (IB: 150 cells/mm^2^ (IQR 130) (**Figure 5**). Lower levels of IB were measured in MASLD/MASH cases at the interface (IB: 80 cells/mm^2^ (IQR 90), p<0.0001). The ICN in the same region was also higher in AIH (390 cells (IQR 580) compared to 140 cells (IQR 220) in MASLD/MASH, p<0.0001).

**Figure 5.**
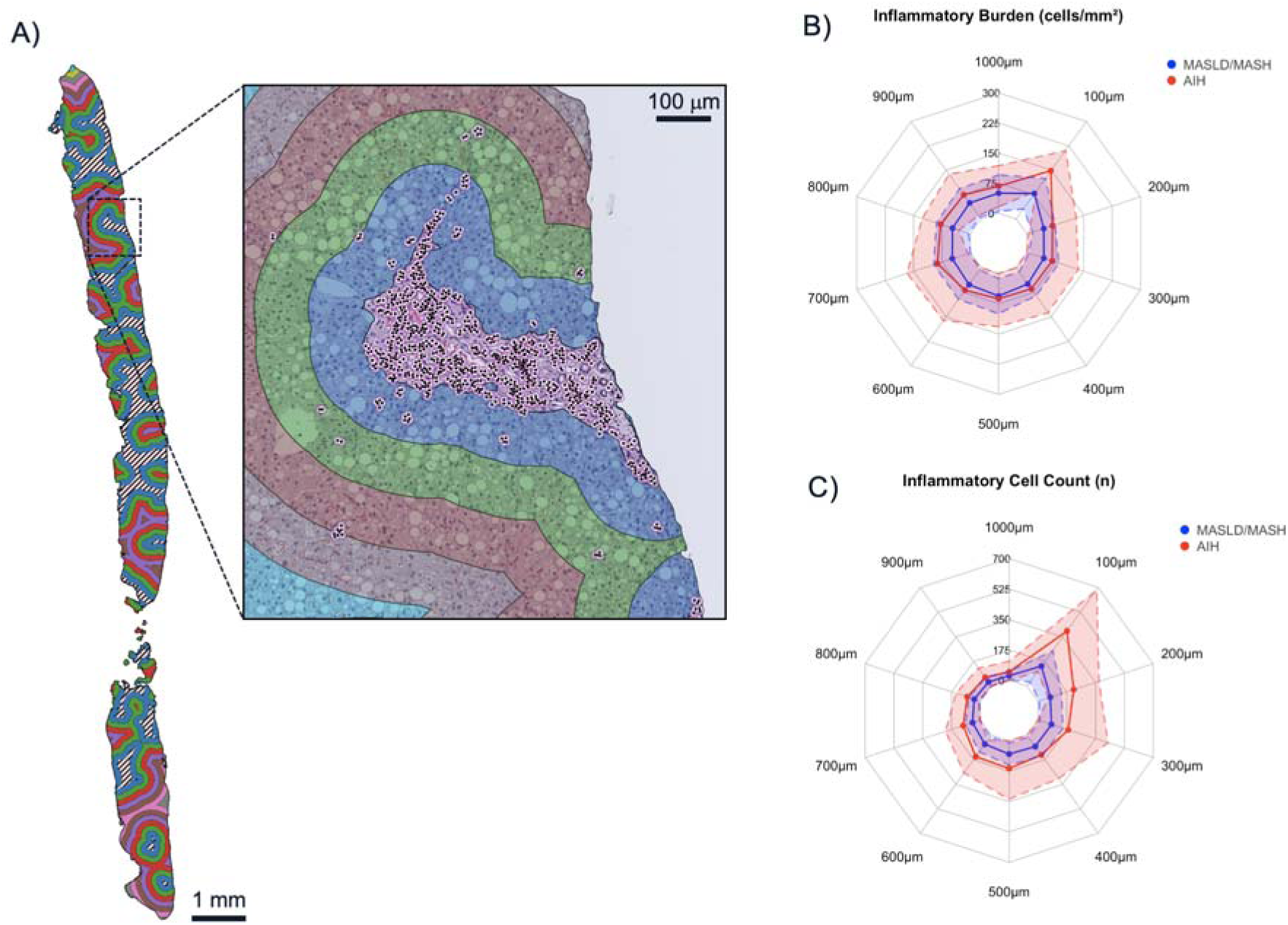
– Regional analysis performed across WSI for inflammatory burden and inflammatory cell numbers in MASLD/MASH (n=327) and AIH (n=63). (A) Inflammatory metrics are extracted from individual regions positioned equidistant from the PT, at intervals of 100μm, in an AIH liver biopsy. (B) Inflammatory burden and (C) inflammatory cell number in MASLD/MASH and AIH, across equidistant regions. Median is represented as a solid line and the upper/lower interquartile range is represented as a dashed line.

In both AIH and MASLD/MASH, the IB was lower at distances of 200–600μm from the PT (2.2-fold lower relative to the interface region, for AIH, and 1.8-fold for MASLD/MASH), but then increased again in lobular regions further away in AIH (700–900μm; 1.3-fold higher relative to the IB at 200μm from PT). In contrast, the ICN decreased gradually across lobular regions in both AIH and MASLD/MASH.

#### Association with scoring of inflammation from manual pathology reads

Grading of inflammation in the MASLD/MASH samples showed good agreement between pathologists for portal tracts and interface region but was less aligned for lobular regions (**Supplementary Figure 4**). Severity of portal inflammation from manual scoring correlated well with ICN count at PT (r_s_=0.46-0.73; p<0.01), but only weakly with IB (r_s_=0.16-0.28; all non-significant). Pathologist scorings for both Ishak and NAS for lobular inflammation correlated weakly with both IB and ICN (r_s_=0.15-0.35; p<0.05 – p<0.34).

Manual grading from single pathologists showed similar trends. For all regions, severity of inflammation from manual pathology correlated better with ICN than with IB (**Figure 6**). In addition, increasing grade of inflammation was more consistent with increasing ICN and IB at the PT and interface regions. Crucially, however, for equivalent grades of portal inflammation the ICN was up to 3.1-fold higher in AIH than in MASLD/MASH and this pattern was seen at portal, interface and lobular regions (**Figure 6**, **Supplementary Table 4**). Of note, ICN at PT correlated well with severity of interface hepatitis in both AIH (r_s_=0.68, p<0.0001) and MASLD/MASH (r_s_=0.56, p<0.0001) (**Supplementary Figure 5**).

**Figure 6.**
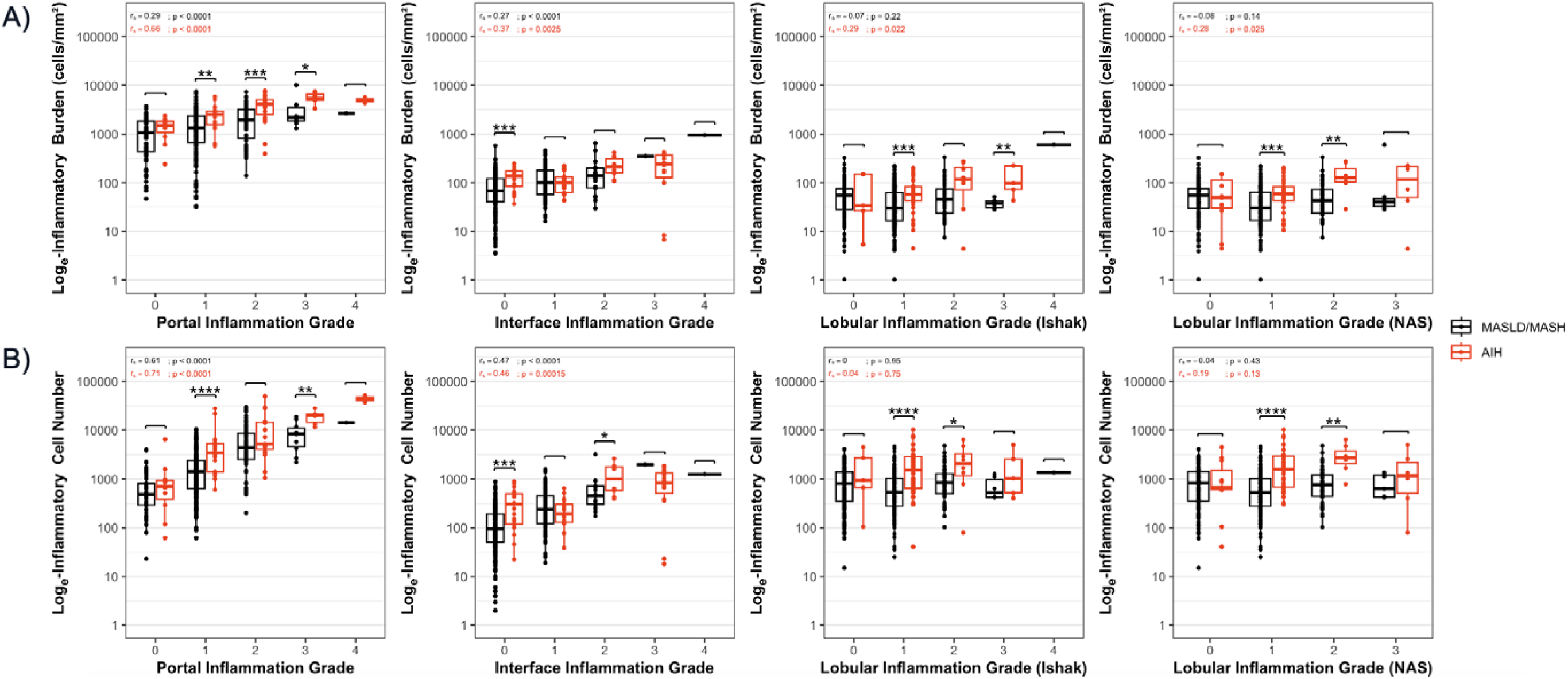
– Inflammation quantified by AI in relation to manual grading by a single pathologist for MASLD/MASH (n=327) and for AIH (n=63). (A) Shows grades relative to IB (B) Shows Ishak grades relative to ICN. Grades shown are based on Ishak scoring at portal and interface regions and by either Ishak or NAS grading at the lobules.

## Discussion

Our aim was to develop an AI solution to identify and quantify PT, steatosis, and inflammation to evaluate 390 MASLD/MASH and AIH cases. The resulting AI system accurately identified and quantified PT and delineated equidistant 100μm regions surrounding them. AI detected more steatosis and larger steatotic vesicles in MASH than in MASLD. Higher levels of inflammation were found across all AIH cases compared to MASLD/MASH cases. Regional distribution of inflammation across regions was similar between MASLD/MASH and AIH and was higher at the PT and interface. However, overall, greater degrees of inflammation were detected in the AIH cases, even for equivalent grades of inflammation from manual pathology. This comprehensive set of detailed metrics can assist pathology readers in differentiating liver disease types, monitoring disease progression and treatment response. The consistent and unbiased assessment offered by such AI/machine learning systems makes them valuable as decision support tools for pathologists.

Manual identification of PT can be time-consuming and challenging due to their variable appearance. This AI system has the potential to speed up this process by directing pathologists’ attention to likely PT. This is particularly relevant for highly fibrotic cases where vascular structures merge together and portal tracts expand resulting in a lack of clear boundary between individual portal tracts^48^. This pipeline could be applied to perform automatic quality control on liver biopsy samples by enumerating total number of PT. Biopsy length is often used as an alternative to PT count for sample adequacy^49^ and this AI system could additionally assist by automatically calculating the biopsy length and the full portal tract per linear cm^22^.

In the NAS scoring system, steatosis scores are performed by assessing the percentage of hepatocytes that contain lipid deposits. Our AI system reports the macrovesicular steatosis fraction (SF) by area and microvesicular steatosis is excluded. Despite this, when comparing the SF to the ratio of macrovesicular steatosis to segmented hepatocytes, the correlation was very strong. This suggests that the SF metric is equivalent to what is graded manually. Our data showed that SF is increased in MASH as compared to MASLD across periportal and lobular regions, however, due to the low number of advanced MASH cases (Fibrosis score of 4) this was not explored^50^. Our system revealed a difference in the median steatotic vesicle size between MASLD and MASH across periportal and lobular areas. Micro– and macrovesicular steatosis exist in both MASLD and MASH, yet there is no clear consensus on whether the pattern of steatotic vesicle size varies between the disease states^51^. These larger droplets in MASH might indicate increased steatogenesis or an increased rate of coalescence, which might be expected to correlate with progressive disease activity in MASH^52^. However, the clinical significance of steatosis is unclear and is not related directly reassociated to mortality or outcomes^53,54^.

Overall, there was broad agreement between AI metrics and pathologist steatosis grading for both MASLD and MASH cases. However, a striking feature of our results is that our SF measurements are under 33%, which does not appear consistent with pathologist steatosis scores greater than 1. Despite this, low SF measurements would match that of low MRI-PDFF that is often recorded^55^. There is evidence of pathologists overestimating steatosis, especially in severe cases^56^ which may contribute to this observed difference. The clinical impact of steatosis not directly associated to mortality or outcomes^53,54^. There is evidence of pathologists overestimating steatosis, especially in severe cases^56^ which may contribute to this observed difference, however a direct comparison of our systems measurements with steatosis grades should be interpreted with caution due to the differences in measurement methods. The clinical impact of steatosis not directly associated to mortality or outcomes^53,54^, however in drug efficacy trials for MASH reduction in liver steatosis can lead to disease activity resolution^53,54^. This AI system could support manual grading of steatosis in such trials.

Overall, our findings show that, as expected, the total level of inflammation is elevated in AIH compared to MASLD/MASH and that AI metrics inflammation correlate with manual grading of inflammation, as long as there is substantial inflammatory cell burden. AI metrics for inflammation (for both ICN and IB) provided more quantitative information of regional distribution than is possible from manual pathology. We found that cases with the same pathologist score may have very different amounts of inflammation depending on the disease aetiology. For example, a portal inflammatory score (Ishak) of 3 or 4 for an AIH case may have considerably higher ICN or IB than a MASLD/MASH case with the same inflammation grade. A pattern of IB was observed in both AIH and MASLD/MASH where inflammation was greater in interface regions and higher 700-900μm from the PT. Similarly, the ICN tended to reduce with increasing distance from the PT with a slight peak of inflammation from 500-600μm from PT. This regional distribution may reflect the biology of the localised inflammatory response. In portal and interface regions, ICN and IB generally increase with higher pathologist scores. In lobular regions there is no correlation, but this may reflect both the notoriously low inter-rater variability for lobular inflammation and the low levels of inflammation found in lobular regions of MASLD/MASH cases^57^.

Although portal inflammation is not typically included classical MASLD/MASH scoring systems such as NAS, it is frequently linked with disease progression^58–61^ and used used to support the disease resolution decisions in MASH trial endpoints. The scoring systems for chronic hepatitis include both portal and lobular scorings in addition to the portal-parenchymal interface. This periportal inflammation correlates well with fibrosis progression in biopsies in chronic hepatitis C^62^ and understanding the potential role of interface hepatitis in MASH progression requires further investigation^59^. A study by Balitzer et al. revealed that without the addition of new stains as well as a change to inflammatory evaluation, the current biopsy methodology for scoring AIH cases led to under-scoring, especially for inflammatory activity^63^; 17% of cases would have been mis-classified as non-AIH. Using our system, the accurate quantification of inflammation at the interface region and graphical identification of PT with higher levels of inflammation will allow pathologists to more easily measure and identify interface hepatitis, avoid misclassification and potentially identify incidental findings.

Further training and test datasets will improve our current system in addition to comparing our system measurements with additional pathologists and scorings. The septa and connective tissue are outlined in highly fibrotic tissue with our AI system, however, this may skew PT enumeration. AI metrics shown to correlate with pathologist grading herein, should nevertheless be interpreted with some caution due to the differences in measurement methods. Furthermore, integration of these tools with further pathological feature detection, such as hepatocyte ballooning, will provide pathologists with vital information. Even though the AIH cases were fewer than the MASLD/MASH cases in this study, we consistently found elevated levels of inflammation in AIH compared to MASLD/MASH. Our AIH cohort was predominantly pediatric (79%) and may exhibit fewer typical disease features than that found in adults^64^. With further dataset training across different populations and multiple clinical trials, our system will be more accurate at spatially analyzing detected features to determine which biological elements of a portal tract and surrounding elements are present in a given region of tissue.

## Conclusion

Portal tract characterization is fundamental for histological scoring and establishing sample adequacy of liver biopsies. This AI system generates detailed visual overlays to assist pathological review, as well as producing automated quantifications of PT, inflammation and steatosis across portal, interface and lobular regions. AI systems show promise and could improve the reproducibility and accuracy of liver biopsy evaluation and consensus decisions in clinical trials. While the use of non-invasive approaches to quantify inflammation are becoming more prevalent, the low levels of IB and ICN that are present in MASLD and MASH will require the high degree of sensitivity as afforded by AI to measure small, anticipated effect sizes that are likely when assessing the impact of anti-inflammatory treatments.

## Acknowledgements

We would like to thank the patients recruited into the studies used in this article for their participation.

Study 1 was registered with registered with clinicaltrials.gov (NCT03551522). Study 2 was registered with UMIN clinical trials registry (UMIN000026145). Study 3 was registered with the ISRCTN registry (ISCRTN39463479) and the National Institute for Health Research portfolio (15912). The collection of study 3 pathology data was funded by Innovate UK (Project number: 101679). Study 4 was registered with clinicaltrials.gov (NCT03198104) and funded by the Eureka Eurostars 2 Grant (E!10124).

## Data availability statement

The portal tract identification system described for the first time in this study is the intellectual property of Perspectum Ltd and the subject of the Patent Cooperation Treaty PCT/IB2025/0530307. Summary data is included in the manuscript or uploaded as online supplemental information. Anonymised individual patient data can be shared upon request or as required by law and/or regulation with qualified external researchers. Approval of such requests is at the discretion of the study sponsors and is dependent on the nature of the request, the merit of the research proposed, the availability of the data, and the intended use of the data.

## Author Contributions

Conceptualization – DW/AM (Equal). Writing (original draft) – DW (Lead), AM (Supporting), SL (Supporting). Data curation – AM (Lead), CB/KHDW/KF/EF/TK/RG (Supporting). Investigation – DW (Lead), AM/CB (Supporting). Formal analysis – AM/CB (Equal). Preparation – DW (Lead), AM/CB/SL (Supporting), Visualisation – AM (Lead), CB/DW (Supporting). Review & Editing – DW/HTB (Equal), AM/HTB/SL/PA/KF/EF/TK/RK/PW/CL/PB/RG (Supporting).

## List of abbreviations

AI: Artificial Intelligence
AIH: Autoimmune Hepatitis
DL: Deep Learning
H&E: Haematoxylin and Eosin
IB: Inflammatory Burden
ICN: Inflammatory Cell Number
SF: Steatosis Fraction
MASLD: Metabolic Dysfunction-associated Steatotic Liver Disease
MASH: Metabolic Dysfunction-associated Steatohepatitis
ML: Machine Learning
MRI: Magnetic Resonance Imaging
NAS: NAFLD Activity Score
PDFF: Proton Density Fat-Fraction
PT: Portal Tract(s)
WSI: Whole Slide Image(s)

