## Supplementary Data for "AI portal tract detection and characterisation for a regional analysis of steatosis and inflammation in MASLD, MASH, and AIH"

*Whole Slide Imaging* H&E-stained liver biopsy slides were digitized to create whole slide images (WSIs) for downstream analysis using Hamamatsu (C12000-02) and Leica Aperio AT2 slide scanners.

*Landmark feature and PT detection* The AI system segments an ensemble of core tissue features in liver histology images (the tissue foreground, regions of connective tissue, bile ducts, hepatic arteries, and portal veins, with the latter three corresponding to the fundamental components of the portal triad). In relation to these segmented features, the AI system performs spatial analysis to count the portal triad elements present within each specific region of connective tissue. A confidence score from 0–3 is assigned to the connective tissue region depending on which of bile duct (+1), portal vein (+1), and hepatic artery (+1) features are present within, or on the tissue region boundary. The score is binary for each feature and additional instances of each within the connective tissue do not increase the score. In line with pathologist-designated annotations of PT, any region of connective tissue with a confidence score of ≥2 was deemed a PT.

*Regional demarcation* The AI system calculates a radial region of 100μm width immediately next to each PT to approximate interface regions. Lobular regions were defined as the difference between the tissue foreground and all candidate PT with a confidence score ≥2. Connective tissue regions with only a single identified triad component were excluded from lobular regions to optimize distinction between portal and lobular inflammatory cells. To probe regional distribution of both inflammation and steatosis, a further set of 9 equidistant 100μm periportal regions extending to a maximum of 1 mm into the liver lobule were generated from the PT boundary (**Figure 1**).

*Steatotic vesicle detection* A bespoke instance segmentation system was trained to detect macrovesicular steatosis (minimum vesicle size >100$\mu m^2$). The training data for this system was generated from a bespoke classical image segmentation pipeline consisting of thresholding and morphological operations for each of the varied slides of our training dataset. Training tiles and masks of these classically segmented images were exported to train a deep learning model for steatosis that could generalise across the highly varied appearance of liver histology slides.

*Inflammatory cell detection* This was based on differences in nucleus size and cell clustering from hepatocytes and other structural cells[1]. In brief, the AI system performed nucleus segmentation using StarDist[2,3], then applied a combination of spatial and morphological filtering to detect the inflammatory subset. Specifically, the algorithm finds dense regions of nuclei whose area is $<50\mu m^2$. These candidate nuclei were then filtered using a nearest-neighbors procedure where the mean of the distance of each candidate nucleus to its 4 nearest neighbors was calculated and nuclei with a mean distance to their nearest neighbors less than a threshold ($10\mu m$) were classified as being inflammatory cell nuclei. Nuclei identified in specific triad structures (e.g. bile ducts) were excluded to remove quantification of confounding closely-packed cholangiocytes and other structural features.

*Feature quantification* Identified features (inflammatory cells, steatotic vesicles, PT, interface and lobular regions, equidistant periportal zones) were represented as abstract geometries whose coordinates were linked using applied geometric and spatial analysis techniques. All regional geometries were used to generate measurements across portal, interface, periportal, and lobular regions within a 1 mm region around every portal tract. Spatial algorithms were implemented in Python using the Geopandas library[4].

*Histological annotation* The AI system was trained and then tested in relation to annotations by the pathologists. The slides were manually annotated for PT using QuPath[5]. Candidate PT annotation required at least two of three characteristic histological components to be visible in the labelled region (hepatic artery, portal vein, bile duct). The final polygon annotations were extracted from each WSI into a GeoJSON file format to train the AI and detect each anatomical feature in the PT.

Annotations of connective tissue, bile ducts, portal veins, and hepatic arteries were collected from 43 individual H&E-stained WSIs from 3 clinical cohorts.
A consensus set of 219 PT annotations from 12 WSI was created by taking the union of all pairwise intersections of PT annotations from three independent pathologists (**Supplementary Table 5**). These consensus annotations formed the ground truth for PT detection in testing of the AI system.
Correlations between measurements were investigated using Spearman's rank correlations (one continuous variable, the other categorical) or Pearson's correlation (both continuous variables) with correlations greater than 0.60 considered strong[6]. Performance of individual pathologists and of AI system in the annotation of PT was evaluated using the F1 score for machine learning evaluation, with F1 scores greater than 0.8 considered good[7]. A modification of this F1 score was also applied, in which each true positive is weighed by the Intersection-over-Union (IoU) of the predicted PT with the ground truth PT (IoU runs between 0-1).

*Landmark feature and portal tract detection* We used 31 mixed liver disease WSIs to train our segmentation models with 12 WSIs used as a held-out test dataset for assessing model performance. We applied a range of augmentations during training, including flips, zooms, rotations, blurring, contrast enhancement, colour augmentation and addition of noise. We trained all models with an 80:20 train:validation split with the Adam optimizer and initial learning rate of $1x10^{-4}$. Image-mask pairs of 256x256 pixels were used for training at a pixel resolution of 0.5 μm/pixel, and the number of un-augmented training tiles are summarized in **Supplementary Table 6**. For each feature we trained a U-net++ semantic segmentation model with an equal weight Dice-BCE loss function for 200 epochs, allowing early stopping after ten epochs of no improvement. Inference was performed on overlapping tiles extracted from WSIs at 0.5 μm/pixel, and tile predictions were represented as geometries, then geometries were combined into slide-level geometries through taking the union of all tile geometries relative to their known position in the slide.
*Lobular region definition* Tissue slicing means that there are genuine PT which will appear with ≤1 triad component in the sample. To minimize bias associated with this effect we excluded connective tissue regions with only a single identified triad component from the lobular region This was done because lobular inflammatory cells

are present at a much lower density than portal inflammatory cells, and therefore lobular measurements of inflammation may be skewed unduly by excluded and inflamed PT.

*Steatotic vesicle detection* Colour thresholding and morphological operations were applied to a subset of 17 WSIs from the study 3 dataset (H&E stain) to generate estimated macro-steatosis segmentations. These segmentations were corrected by a pathologist (TK) using the open-source software QuPath[5]. Twelve annotated WSIs were divided into overlapping tiles (512x512px) and used to train a U-Net machine learning model to segment steatosis, and the remainder was used for cross-validation[8]. The network achieved a Sorensen-Dice precision of 0.95 and recall of 0.91 on the validation tiles. All MASLD/MASH cases (n=327) were subsequently analyzed using the trained U-net to generate steatosis segmentations and proportionate area.

*Inflammatory cell detection* Inflammation detection was completed in a 2-step process: nuclei segmentation and nuclei classification. A pretrained Stardist model was used to segment the nuclei in the WSI with the centroid location and size of each being recorded[2,3]. The mean of the distance of each nucleus to its neighbors was calculated and nuclei with a small mean distance to their nearest neighbors were classified as inflamed. Morphological dilation was then applied to the binary mask of the inflamed nuclei to get a mask of the inflamed regions, an example of which is shown in Figure 2. The IB was calculated by dividing the sum of the inflamed-nuclei mask by the sum of the foreground mask. Inflammatory cell identification was validated by a process of manual review by all 4 pathologists.

Supplementary Figure 1: Study design and populations

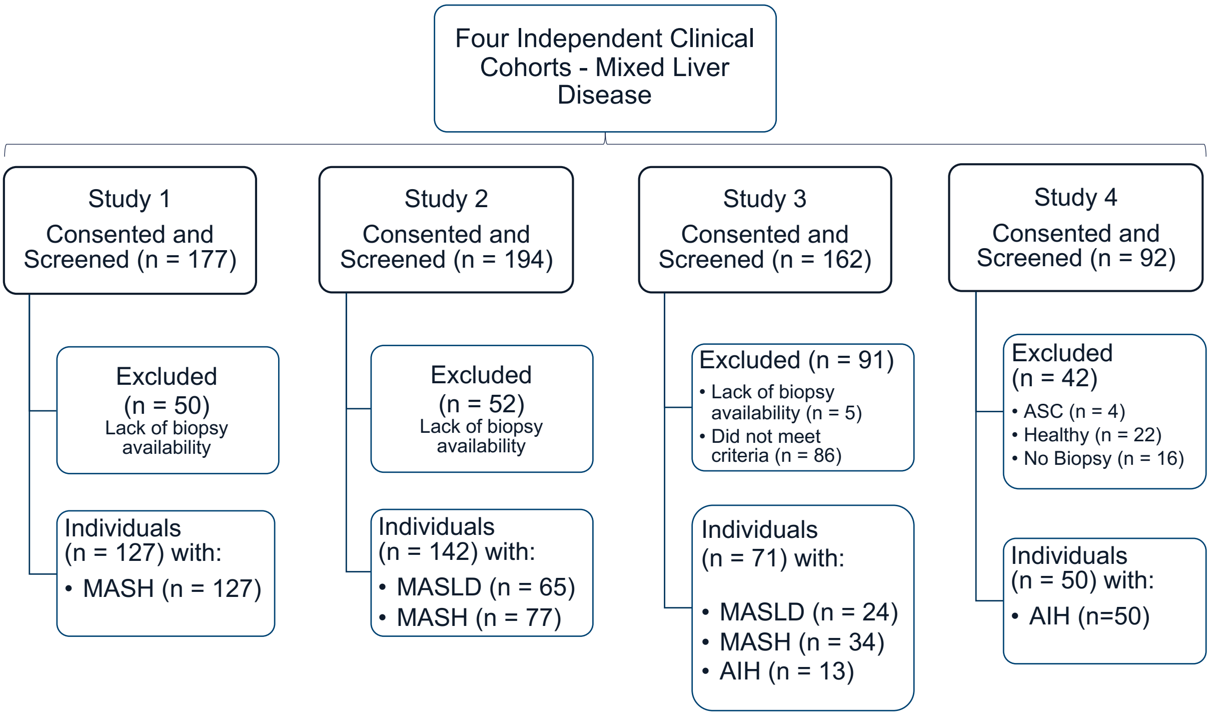

Supplementary Figure 2 – Detection of PT along length of biopsy sample (A) and correlation of total PT with length of sample (B).

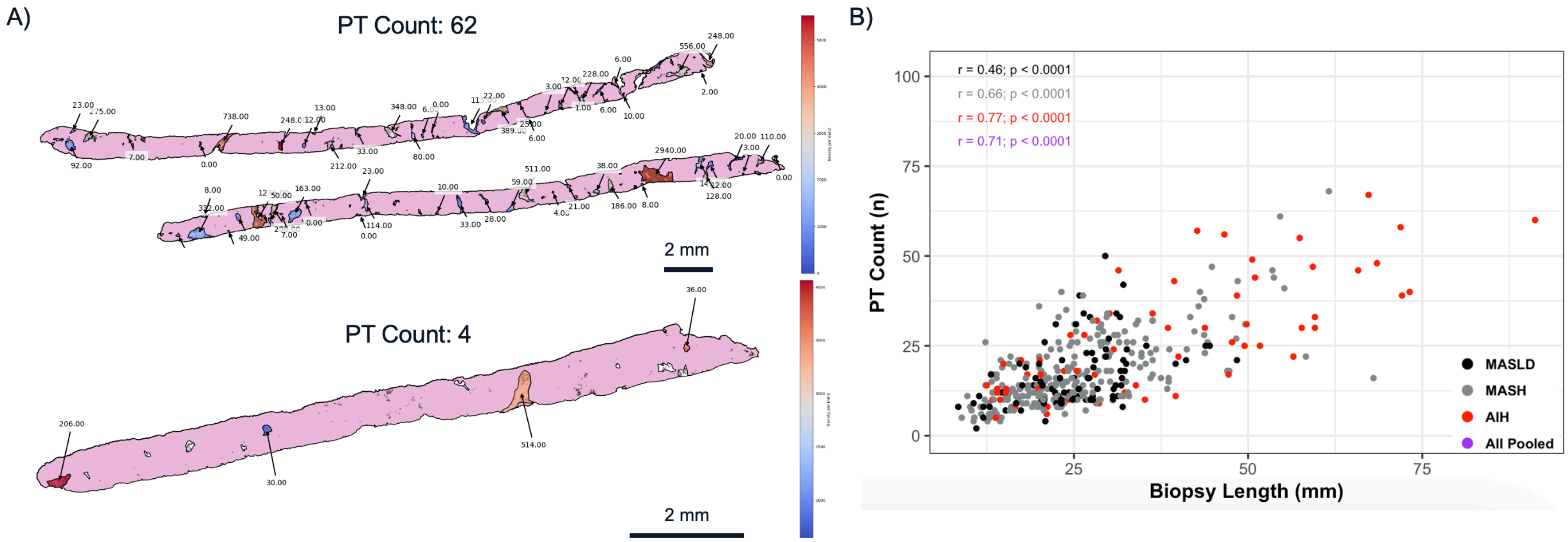

Supplementary Figure 3 – AI derived metrics in samples with different grade of steatosis from manual reads (NAS). Steatosis fraction (A) or vesicle to hepatocyte fraction (C) was compared to each individual pathologist's grading in MASLD (n = 18) and MASH (n = 34). Steatosis fraction (B) or vesicle to hepatocyte fraction (D) was compared a single pathologist grading in MASLD (n = 89) and MASH (n = 238). Correlation of steatosis fraction to vesicle to hepatocyte fraction in all 327 samples (E).

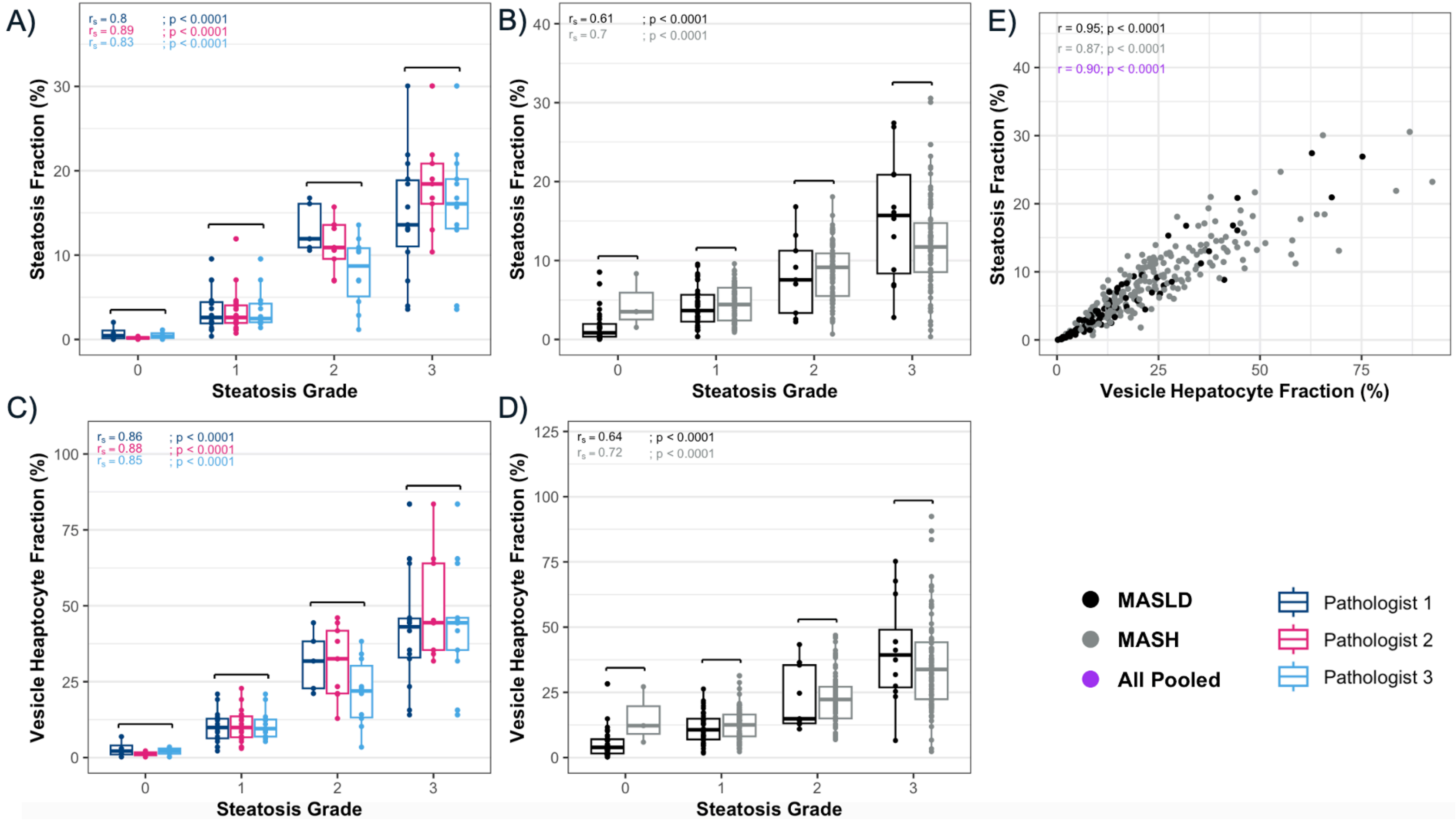

Supplementary Figure 4 – Correlations between AI derived metrics and the pathologist scorings for inflammatory burden (A) and inflammatory cell number (B) in 52 MASLD/MASH slides.

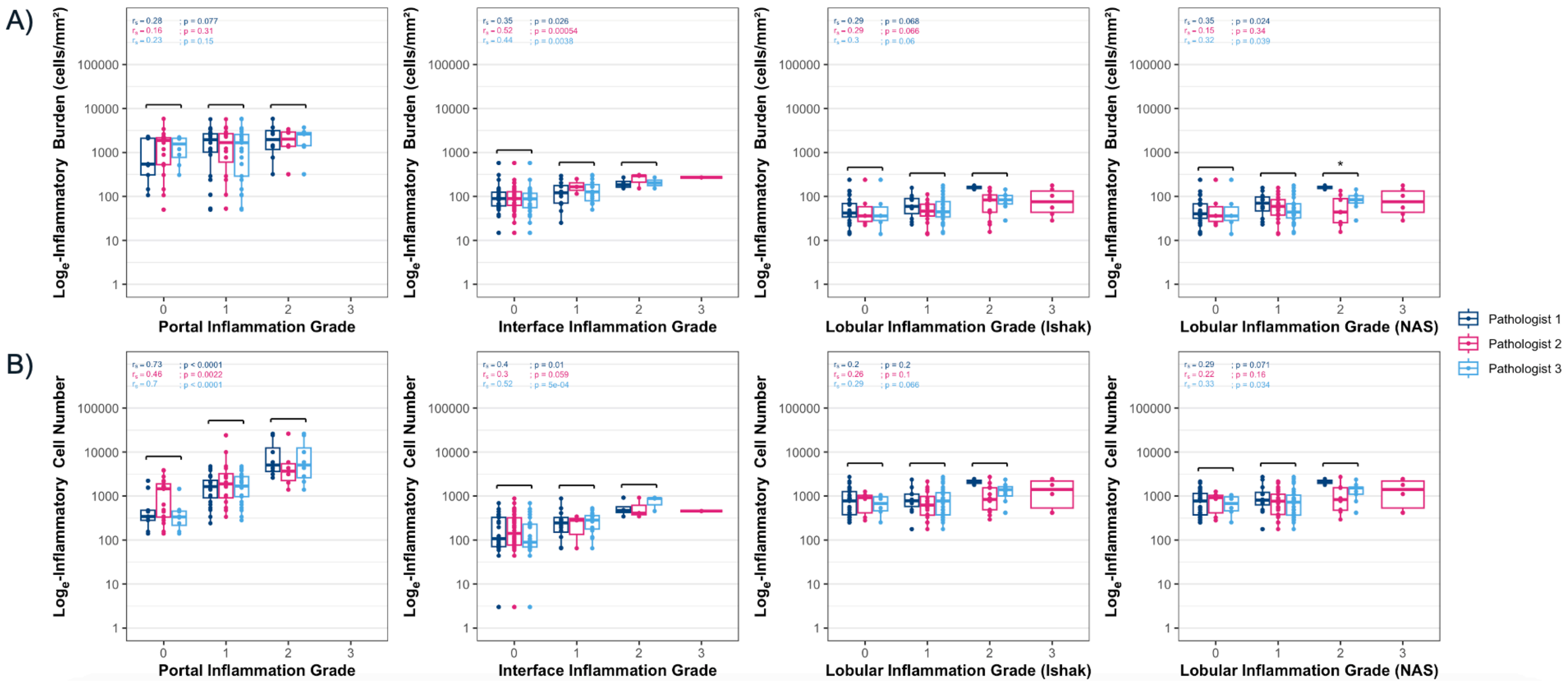

Supplementary Figure 5 – Inflammation at portal tract compared with grading of interface hepatitis, for MASLD/MASH (n = 258) and AIH (n = 63). (A) IB and (B) ICN.

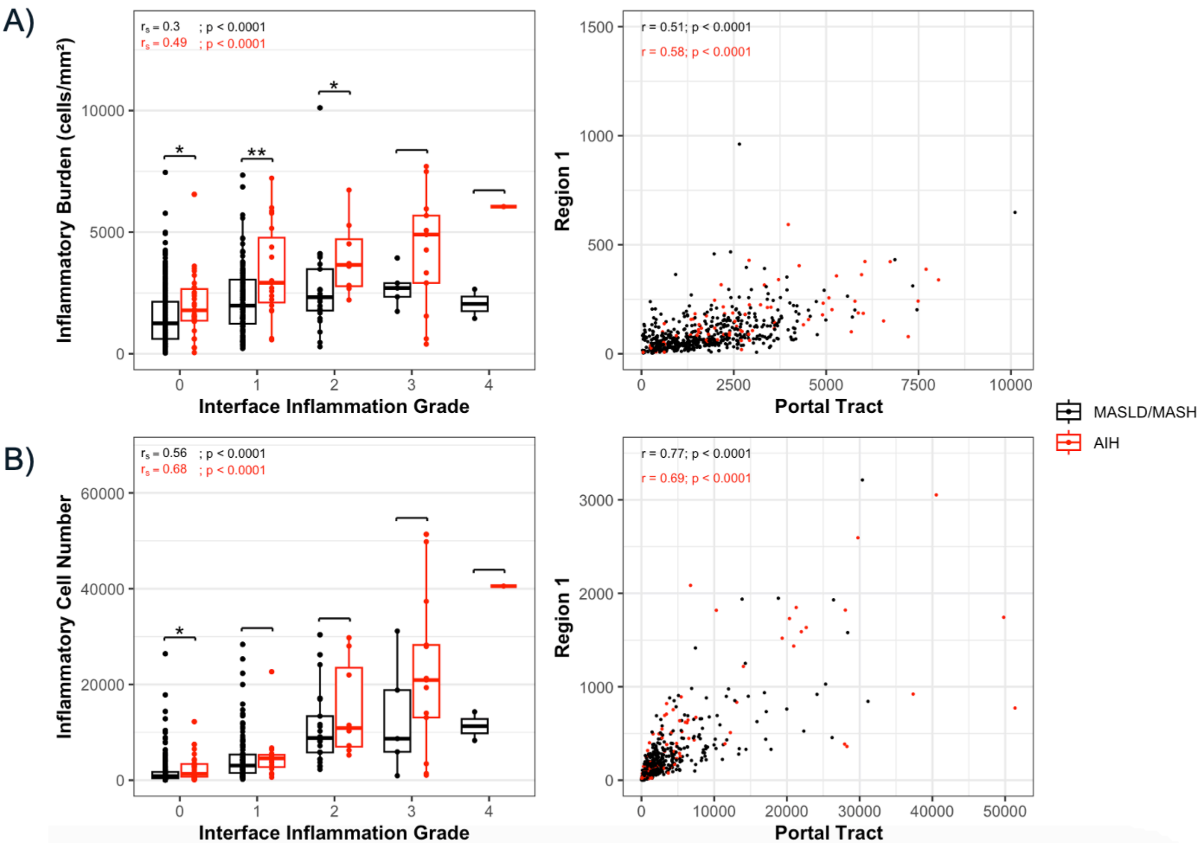

Supplementary Table 1: Study populations

*Shown as n (%). #Shown as Mean± SD. §Shown as median (IQR).

| | **All (n=390)** | **MASLD (n=89)** | **MASH (n=238)** | **AIH (n=63)** |
|---|---|---|---|---|
| **Demographics#** | | | | |
| Age (yr) | 51 ±19 | 55 ±15 | 56 ±12 | 21 ±17 |
| BMI (kg/m$^2$) | 29.5 ±6.8 Unknown (n=69) | 27.9 ±5.1 Unknown (n=3) | 32.6 ±5.7 Unknown (n=62) | 22.6 ±6.1 Unknown (n=4) |
| **Sex*** | | | | |
| Male | 186 (48%) | 61 (69%) | 99 (42%) | 26 (41%) |
| Female | 201 (52%) | 28 (21%) | 139 (58%) | 34 (54%) |
| Not specified | 3 (<1%) | - | - | 3 (5%) |
| Hypertension | 68 (17%) | 27 (30%) | 41 (17%) | BP <133/85mmHg |
| **Treatment*** | | | | |
| Yes - Steroids/ Azathioprinum | 35 (56%) | - | - | 35 (56%) |
| No | 15 (24%) | - | - | 15 (24%) |
| Unknown | 13 (20%) | - | - | 13 (20%) |
| **Biopsy sampling and digitization#** | | | | |
| No. of biopsy samples and WSI | 390 | 89 | 238 | 63 |
| Time between clinical visit and biopsy (d) | 3 (n=50) | - | - | 3 (n=50) |
| Length of biopsy (mm) | 22 ±9 (n=108) | 25 ±9 (n=34) | 26 ±5 (n=24) | 20±9 (n=50) |
| **Manual pathology scoring#** | | | | |
| Number of PT per biopsy | 19 ±12 (n=108) | 12 ±2 (n=34) | 11± 4 (n=24) | 29 ±17 (n=50) |

| | All (n=390) | MASLD (n=89) | MASH (n=238) | AIH (n=63) |
|---|---|---|---|---|
| PT per length of sample (/10mm) | 7 ±3 (n=108) | 5 ±1 (n=34) | 5 ±1 (n=24) | 7 ±3 (n=50) |
| **Interface Inflammation Score*** | | | | |
| Grade 0 | 238 (61%) | 66 (74%) | 151 (63%) | 21 (33%) |
| Grade 1 | 106 (27%) | 20 (23%) | 69 (29%) | 17 (27%) |
| Grade 2 | 28 (7%) | 2 (2%) | 16 (7%) | 10 (16%) |
| Grade 3 | 16 (4%) | - | 1 (1%) | 15 (24%) |
| Grade 4 | 1 (1%) | 1 (1%) | - | - |
| **Lobular Inflammation* (Ishak)** | | | | |
| Grade 0 | 117 (30%) | 57 (64%) | 55 (23%) | 5 (8%) |
| Grade 1 | 207 (53%) | 27 (30%) | 135 (57%) | 45 (71%) |
| Grade 2 | 54 (14%) | 4 (5%) | 42 (18%) | 8 (13%) |
| Grade 3 | 11 (3%) | - | 6 (2%) | 5 (8%) |
| Grade 4 | 1 (<1%) | 1 (1%) | - | - |
| **Lobular Inflammation* (NAS)** | | | | |
| Grade 0 | 122 (31%) | 56 (63%) | 55 (23%) | 11 (17%) |
| Grade 1 | 202 (52%) | 28 (32%) | 135 (57%) | 39 (62%) |
| Grade 2 | 53 (14%) | 4 (4%) | 42 (18%) | 7 (11%) |
| Grade 3 | 13 (3%) | 1 (1%) | 6 (2%) | 6 (10%) |
| Grade 4 | - | - | - | - |
| **Portal Inflammation Score* (Ishak)** | | | | |
| Grade 0 | 81 (21%) | 28 (32%) | 42 (18%) | 11 (17%) |
| Grade 1 | 200 (51%) | 43 (48%) | 132 (55%) | 25 (40%) |
| Grade 2 | 90 (23%) | 17 (19%) | 54 (23%) | 19 (30%) |
| Grade 3 | 16 (4%) | - | 10 (4%) | 6 (10%) |
| Grade 4 | 3 (1%) | 1 (1%) | - | 2 (3%) |
| **Fibrosis Grade* (NAS)** | | | | |

| | All (n=390) | MASLD (n=89) | MASH (n=238) | AIH (n=63) |
|---|---|---|---|---|
| Grade 0 | 78 (20%) | 21 (24%) | 4 (2%) | 53 (85%) |
| Grade 1 | 56 (14%) | 17 (19%) | 37 (15%) | 2 (3%) |
| Grade 2 | 74 (19%) | 13 (14%) | 61 (26%) | - |
| Grade 3 | 130 (34%) | 24 (27%) | 102 (43%) | 4 (6%) |
| Grade 4 | 51 (13%) | 14 (16%) | 33 (14%) | 4 (6%) |
| **Steatosis Grade* (NAS)** | | | | |
| Grade 0 | 91 (23%) | 30 (34%) | 3 (1%) | 58 (92%) |
| Grade 1 | 121 (31%) | 38 (43%) | 79 (33%) | 4 (6%) |
| Grade 2 | 90 (23%) | 9 (10%) | 80 (34%) | 1 (2%) |
| Grade 3 | 88 (23%) | 12 (13%) | 76 (32%) | - |

Supplementary Table 2: AI metric overview

| **AI Derived Metrics§** | | | | | | |
|---|---|---|---|---|---|---|
| | All (n=390) | MASLD (n=89) | MASH (n=238) | AIH (n=63) | p-value MASLD vs MASH | p-value MASLD/ MASH vs AIH |
| Length of biopsy (mm) | 27.2 (12.6) | 25.7 (9.9) | 23.4 (12) | 32.3 (29.6) | NS | p < 0.01 |
| Number of PT per biopsy | 19 (12) | 14 (11) | 15 (12) | 22 (24) | NS | N/A |
| PT per length of sample (/10mm) | 7 (3) | 6 (4) | 6 (4) | 7 (41) | NS | N/A |
| SF | 7% (3) | 3% (6) | 8% (7) | N/A | p < 0.0001 | N/A |

| AI Derived Metrics§ | | | | | | |
|---|---|---|---|---|---|---|
| | All (n=390) | MASLD (n=89) | MASH (n=238) | AIH (n=63) | p-value MASLD vs MASH | p-value MASLD/ MASH vs AIH |
| Total ICN | 3200 (4700) | 3400 (4000) | 2400 (3800) | 6400 (12400) | $p < 0.05$ | $p < 0.0001$ |
| IB (cells/mm$^2$) – Interface | 87 (106) | 100 (90) | 70 (90) | 150 (130) | $p < 0.01$ | $p < 0.0001$ |
| IB (cells/mm$^2$) – Portal | 1700 (2000) | 1400 (2200) | 1400 (1700) | 2700 (2600) | NS | $p < 0.0001$ |
| IB (cells/mm$^2$) – Lobular | 40 (50) | 50 (40) | 30 (50) | 60 (80) | $p < 0.01$ | $p < 0.001$ |
| ICN (n) – Interface | 170 (270) | 180 (240) | 120 (210) | 390 (580) | $p < 0.05$ | $p < 0.0001$ |
| ICN (n) – Portal | 1700 (3700) | 1900 (2800) | 1400 (2500) | 4600 (9500) | NS | $p < 0.0001$ |
| ICN (n) – Lobular | 750 (1100) | 800 (900) | 580 (900) | 1500 (2200) | NS | $p < 0.0001$ |

Supplementary Table 3. Performance of individual pathologists and of AI model in the annotation of PT. AI model performance was compared to a consensus set of 219 PT defined from the unions between individual pathologist annotations.

| | Anatomical annotation of PT* | | | |
|---|---|---|---|---|
| | Inter-operator performance for manual annotation | | | Performance of AI model vs manual consensus |
| Mean Value ±SD | Pathologist 1 vs 2 | Pathologist 2 vs 3 | Pathologist 3 vs 1 | AI vs consensus annotation |
| F1 Score | 0.68 ± 0.25 | 0.58 ± 0.16 | 0.72 ± 0.11 | 0.66 ± 0.18 |
| Modified F1 Score | 0.54 ± 0.23 | 0.42 ± 0.14 | 0.60 ± 0.14 | 0.54 ± 0.16 |
| Precision | 0.64 ± 0.24 | 0.83 ± 0.11 | 0.61 ± 0.14 | 0.70 ± 0.15 |
| Recall | 0.75 ± 0.28 | 0.47 ± 0.16 | 0.92 ± 0.09 | 0.67 ± 0.24 |

* As Mean Value ±SD

Supplementary Table 4. Regional ICN at each grade of inflammation from manual histology.

| Inflammation Grade (Ishak) | MASLD (n=89) | MASH (n=238) | AIH (N=63) |
|---|---|---|---|
| | ICN | ICN | ICN |
| Portal | | | |
| 0 | 540 ±860 | 480 ±480 | 700 ± 530 |
| 1 | 2200 ±2200 | 1000 ±1500 | 3500 ± 3900**** |
| 2 | 5300 ±8300 | 4400 ±5700 | 5200 ± 10400 |
| 3 | NA | 8300 ±6312 | 20100 ± 6500** |
| 4 | 14300 | NA | 44400 ± 7000 |
| Interface | | | |
| 0 | 160 ±180 | 80 ±120 | 310 ±380*** |
| 1 | 320 ±470 | 210 ±310 | 200 ±310 |
| 2 | 560 ±100 | 400 ±480 | 1100 ±1200* |
| 3 | NA | 1900 | 830 ±800 |
| 4 | 1250 | NA | NA |
| Lobular | | | |
| 0 | 800 ±830 | 780 ±1200 | 940 ± 2000 |
| 1 | 700 ±960 | 500 ±760 | 1500 ± 2200**** |
| 2 | 600 ±730 | 840 ±810 | 2100 ± 2200* |
| 3 | NA | 540 ±570 | 1000 ± 2000 |
| 4 | 1400 | NA | NA |

Supplementary Table 5. WSI scoring between pathologists

| | Pathologist 1 | Pathologist 2 | Pathologist 3 | Pathologist 4 |
|---|---|---|---|---|
| **AIH** WSI | | | | |
| PT detection to test AI model | N=0 | N=0 | N=0 | N=0 |
| Ishak for inflammation | N=0 | N=0 | N=0 | N=63 |
| **MASLD** WSI | | | | |
| PT detection to test AI model | N=8 | N=8 | N=8 | N=8 |
| Ishak for inflammation | N=18 | N=18 | N=89 | N=0 |
| NAS for inflammation | N=18 | N=18 | N=89 | N=0 |
| NAS for steatosis | N=18 | N=18 | N=89 | N=0 |
| **MASH** WSI | | | | |
| PT detection to test AI model | N=4 | N=4 | N=4 | N=4 |
| Ishak for inflammation | N=34 | N=34 | N=238 | N=0 |
| NAS for inflammation | N=34 | N=34 | N=238 | N=0 |
| NAS for steatosis | N=34 | N=34 | N=238 | N=0 |

Supplementary Table 6. Training tile numbers

| Feature | No. Images | No. WSI |
|---|---|---|
| Bile duct | 1167 | 21 |
| Steatotic vesicles | 34901 | 30 |
| Hepatic artery | 1288 | 31 |
| Portal vein | 1042 | 16 |
| Connective tissue | 7220 | 19 |
| Tissue foreground | 9730 | 11 |
